# Effect of diabetic kidney disease complications on fatigability in patients with type 2 diabetes mellitus

**DOI:** 10.1101/2024.04.27.24306277

**Authors:** Yuma Hirano, Daisuke Tsuriya, Kenichi Kono, Tomoyuki Fujikura, Tomoya Yamaguchi, Kento Matsushita, Yurina Yokoyama, Katsuya Yamauchi, Yusuke Nishida

**Author notes:** **Corresponding author:** (YH).

## Abstract

**Purpose:** Type 2 diabetes mellitus (T2DM) emphasizes the maintenance of high levels of physical activity and requires interventions tailored to the characteristics of each patient. We hypothesized that T2DM combined with diabetic kidney disease (DKD) could increase skeletal muscle fatigability, becoming a specific contributor to physical inactivity. This study aimed to determine the effects of DKD complications on fatigability and the relationship between fatigability and physical activity in patients with T2DM.

**Methods:** The participants were 50 patients with T2DM aged 40–65 years with an estimated glomerular filtration rate of 30 ml/min/1.73 m2 or higher. An experimental protocol was performed using a isokinetic dynamometer to assess muscle function. Fatigability (maximal voluntary concentric contraction [ΔMVCC] velocity, maximal voluntary isometric contraction [ΔMVIC] torque) was calculated, indicating the decrease in angular velocity and muscle strength associated with the exercise task. The patient characteristics, physical activity (IPAQ-SV), knee extension strength, and skeletal muscle index were evaluated. Participants were divided into two groups (DKD and non-DKD) according to the presence or absence of DKD, and ΔMVCC velocity and ΔMVIC torque were compared.

**Results:** ΔMVCC velocity was significantly higher in the DKD group than that in the non-DKD group (p<0.05). Similarly, ΔMVIC torque was significantly higher in the DKD group compared with the non-DKD group (p<0.05). Subgroup analysis showed that ΔMVCC velocity was independently associated with physical activity in the DKD group (odds ratio: 0.045, 95% confidence interval: 0.913–0.999).

**Conclusion:** Fatigability increased with DKD in patients with T2DM and may be related to physical activity.

## Introduction

Type 2 diabetes mellitus (T2DM) is an important chronic disease characterized by chronically elevated blood glucose levels. Maintaining a high level of physical activity is crucial for controlling blood glucose levels and preventing complications. Understanding the mechanism of reduced physical activity in patients with T2DM, with its diverse pathological conditions due to complications, poses a challenge. Therefore, clarifying the mechanism of reduced physical activity specific to each patient’s complications is essential. In this study, we focused on elucidating the factors associated with the amount of physical activity in patients with T2DM and renal dysfunction, specifically those with diabetic kidney disease (DKD). DKD accounts for >40% of the underlying diseases in patients undergoing dialysis in Japan, emphasizing a significant need for intervention.

T2DM is commonly associated with several musculoskeletal disorders [1] that pose significant barriers to regular exercise and physical activity [2,3]. Musculoskeletal disorders involve decreased muscle strength and power and increased muscle fatigability [4]. In contrast to chronic fatigue, fatigability is conceptualized as a symptom associated with physical activity and defined as a reversible, short-term, activity-related decrease in muscle strength, power, or speed [5]. Fatigue symptoms may be chronic, whereas fatigability is reversible with rest [6]. Fatigability is associated with decreased exercise tolerance and adherence [7]. Additionally, the ability to perform repetitive or prolonged daily physical activities, particularly walking, climbing stairs, and carrying loads, is limited by fatigability [8].

Previous reports have shown that patients with prediabetes and T2DM had greater fatigability than did healthy individuals [4,9,10]. Fatigability is associated with skeletal muscle atrophy, mitochondrial dysfunction, decreased hemoglobin concentration, and decreased muscle blood flow [11]. Among these, skeletal muscle atrophy and reduced muscle blood flow are consistent with the characteristics of patients with chronic kidney disease (CKD), including mild cases [12–15]. Therefore, fatigability in T2DM may increases with DKD, potentially hindering physical activity.

However, few studies have examined the effects of DKD on fatigability in patients with T2DM. Additionally, none have examined the relationship between fatigability and physical activity. Therefore, we hypothesized that T2DM combined with DKD would be associated with greater fatigability and reduced physical activity compared with T2DM alone. In this study, we compared fatigability in patients with T2DM and DKD and evaluated the effects of comorbid DKD on fatigability. We also assessed the relationship between fatigability and physical activity, aiming to identify factors associated with physical activity levels in patients with DKD and inform the planning of personalized exercise programs.

## Material and methods

### Study design, setting, participants, and addressing bias

This was a single-center, cross-sectional study of patients with T2DM who visited outpatient clinics between April 2020 and March 2023. The inclusion criteria were: (a) age between 40 and 65 years, (b) ambulatory independence, and (c) estimated glomerular filtration rate (e-GFR) of 30 ml/min/m2 or higher. The exclusion criteria were: (a) inability to provide consent to participate in the study, (b) coexisting severe cardiac or pulmonary disease, and (c) inability to complete the study protocol due to orthopedic or psychiatric diseases. The ultimate goal of this study was to prevent poor glycemic control and progression of renal complications due to physical activity restriction. Therefore, focusing on patients with early DKD was necessary; we enrolled patients with e-GFR ≥30 mL/min/1.73 m^2^. Furthermore, a significant difference in fatigability between younger and older patients has been reported [16]. Therefore, to minimize the influence of age, only patients aged 40–65 years were included in this study. Written informed consent was obtained from all participants before they participated in the study. The study was approved by the Clinical Research Ethics Committee of the Hamamatsu University School of Medicine (20-002). This cross-sectional study followed the Strengthening the Reporting of Observational Studies in Epidemiology guidelines for reporting cross-sectional studies.

### Outcomes

Our main outcomes were fatigability and physical activity.

Fatigability was assessed by an experimental protocol using an isokinetic dynamometer for muscle function assessment (SAKAImed, BIODEX System3), according to previous studies [16] (Fig 1). The experimental protocol consisted of baseline and dynamic fatiguing tasks (DFT). The participants were seated on an isokinetic dynamometer for muscle function assessment, with their hip joints flexed at 90° and their shoulders, hips, and ankles immobilized. The exercises performed in this study included maximal voluntary concentric contractions (MVCC) and maximal voluntary isometric contractions (MVIC) during knee extension. Measurements were taken regarding angular velocity (deg/s) for the MVCC and torque (Nm) for the MVIC. The participants were verbally instructed to use maximum velocity in all MVCCs and maximum force in all MVICs. During each cycle in the MVCC, the participants performed knee extension by contracting the muscles as fast as possible in a 90° range of motion. After the contraction was complete, the muscles were relaxed and passively returned to the starting position by the machine. The experimental protocol was performed by three individuals, one physician, and two physiotherapists trained in measurement. At baseline, the participants performed three MVICs with a rest between each trial. Fifteen MVCC cycles were performed at a load of 20% of the maximum torque value measured by the MVIC. At each MVIC, maximal muscle force was exerted for 5 s. The DFT consisted of 30 consecutive MVCCs once every 3 s, followed immediately by the MVIC. This was considered a set, and three sets were performed consecutively. The maximum value of the three MVICs measured in the baseline task was taken as the base MVIC, and the average value of the fastest five consecutive times of the 15 MVCCs was taken as the base MVCC. Additionally, for each set of MVCCs in the DFT, the average value (v1 to v3) of the last 5 out of 30 contractions was calculated. From base MVCC and base MVIC, the rate of decrease in the third set of MVCC and MVIC in DFT was quantified as ΔMVCC velocity and ΔMVIC torque, respectively, and this was defined as fatigability.

**Fig 1.**
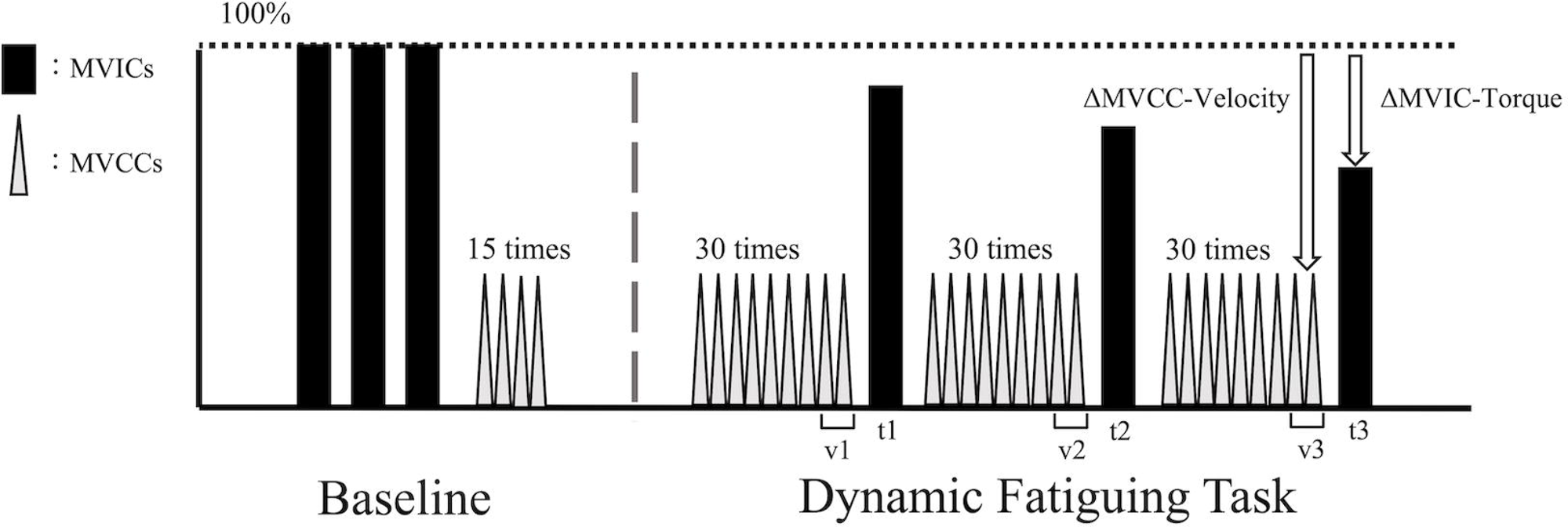
Experimental protocol for the assessment of fatigability. Fatigability was assessed by performing an experimental protocol using an isokinetic dynamometer to assess muscle function. The exercises performed in this study included maximal voluntary concentric contractions (MVCC) and maximal voluntary isometric contractions (MVIC) during knee extension. The black and gray bars represented MVIC and MVCC, respectively. At baseline, the participants performed three MVICs. Subsequently, they performed 15 MVCCs at a load of 20% of the maximum torque value measured by the MVIC. The dynamic fatiguing task consisted of 30 consecutive MVCC cycles, one every 3 s, followed by MVIC (t1∼t3) performed immediately after each MVCC cycle. This was performed in one set and continued for three sets.

Physical activity was assessed using the Japanese Short Version of the International Physical Activity Questionnaire (IPAQ). The questionnaire assessed the amount of physical activity in each category (high intensity, moderate intensity, and walking) over 7 days. Metabolic equivalents (METs) of 8.0, 4.0, and 3.3 were assigned to vigorous-intensity physical activity, moderate-intensity physical activity, and walking, respectively, according to official IPAQ guidelines [17]. The METs were multiplied by the activity time of each item and the number of performance days per week, and the amount of physical activity for each item and its total value were calculated as the amount of physical activity (METs min/week). Moreover, the participants were divided into three groups according to the official IPAQ guidelines: low physical activity (<600 METs min/week), moderate physical activity (600–2999 METs min/week), and high physical activity (>3000 METs min/week).

### Exposures

The primary exposure variable was the presence or absence of DKD. The participants were divided into DKD and non-DKD groups based on the presence or absence of DKD. DKD was defined as albuminuria ≥30 mg/gCr or renal dysfunction without albuminuria (eGFR <60 ml/min/1.73 m2). The eGFR was calculated according to the Japanese estimation formula using serum creatinine concentration [18].

### Potential modifiers

Knee extension strength and skeletal muscle mass index (SMI) were assessed as potential confounders of fatigability and physical activity levels. Knee extension strength and skeletal muscle mass are known functions of the musculoskeletal system. Reports suggest their tendency to decrease with renal impairment and may be related to fatigue [11]. Additionally, they are associated with the amount of daily physical activity in patients with CKD [19]. For knee extension strength, we used the maximum of the three MVICs measured at the baseline of the experimental protocol divided by body weight. Skeletal muscle index was assessed using a body composition analyzer (InBody Japan, InBody 270). The participants stood at the machine after entering personal information such as height, age, and sex. The participants were assessed in a static standing position.

### Covariates

The other variables of interest included age and sex as demographic characteristics. Fatigability increases with age [16, 20], and men are more likely to have increased fatigability than women [21].

### Statistical analysis

The participant characteristics are reported as mean (standard deviation [SD]) or median (interquartile range [IQR]) for continuous variables and as counts and proportions for categorical variables. Inferential statistical analyses using t-tests, chi-square, and Mann-Whitney U tests were performed to compare the characteristics of clinical disease information, patient demographics, fatigability, knee extension strength, and SMI between patients with and without kidney disease. Comparisons of fatigability between patients with and without kidney disease were performed using analysis of covariance, including age, sex, knee extensor strength, and SMI as adjustment variables. Moreover, ordinal logistic regression analysis examined the association between physical activity and fatigability in all participants, including subgroup analysis for the DKD and non-DKD groups. Independent variables included fatigability, age, sex, knee extensor strength, and SMI. Statistical significance was set at p < 0.05. IBM SPSS version 26 was used for data analysis.

## Results

### Participant characteristics

Of the 73 patients who met the selection criteria, 50 were included in the analysis, and 23 were excluded. After classification, 31 and 19 patients were assigned to the DKD and non-DKD groups, respectively (Fig 2). All 50 participants had complete data for analysis. The participants had a median age of 54.5 (IQR, 48.3–61.8) years, a mean BMI of 27.5 ± 5.3 kg/m2, and 52.0% were male individuals. Among the participants, 56.0% had hypertension, 68.0% had dyslipidemia, and 20.0% had coexisting cardiovascular disease (Table 1).

**Fig 2.**
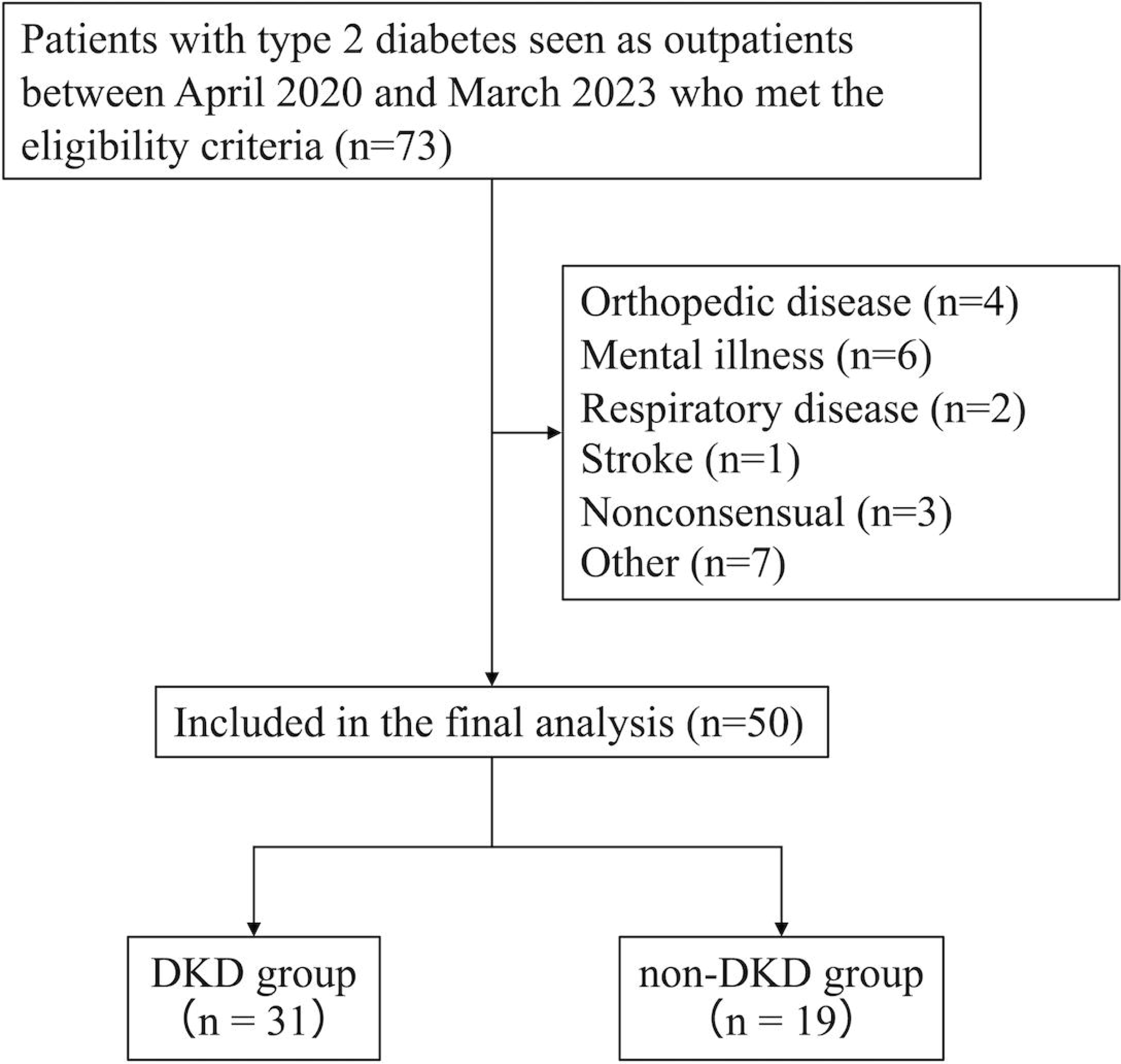
Selection of participants for analysis and assignment to the two groups. Of the 73 patients who met the selection criteria, 50 were included in the final analysis, and 23 were excluded. After classification, 31 and 19 patients were assigned to DKD and non-DKD groups, respectively. *DKD* diabetic kidney disease.

**Table 1.** Characteristics of the study participants.

|  | All<br>(N = 50) | DKD group<br>(n = 31) | Non-DKD group<br>(n = 19) | *p*-Value |
| --- | --- | --- | --- | --- |
| Age (years) | 54.5 (48.3–61.8) | 57.0 (50.0–62.0) | 51.0 (48.0–59.5) | 0.347 |
| Male [N, (%)] | 26 (52.0) | 15 (48.4) | 11 (57.9) | 0.514 |
| BMI (kg/m<sup>2</sup>) | 27.5 ± 5.3 | 27.1 ± 5.4 | 28.6 ± 5.0 | 0.491 |
| Current smoking [N, (%)] | 9 (18.0) | 4 (12.9) | 5 (26.3) | 0.205 |
| Diabetic retinopathy [N, (%)] | 8 (16.0) | 7 (22.6) | 1 (5.3) | 0.108 |
| Hypertension [N, (%)] | 28 (56.0) | 18 (58.1) | 10 (52.6) | 0.707 |
| Hyperlipemia [N, (%)] | 34 (68.0) | 20 (64.5) | 14 (73.7) | 0.500 |
| Cardiovascular diseases [N, (%)] | 10 (20.0) | 5 (16.1) | 5 (26.3) | 0.301 |
| HbA1c (%) | 7.6 (7.1–8.2) | 7.6 (7.1–8.2) | 7.5 (7.0–8.1) | 0.719 |
| Hemoglobin (g/dl) | 14.7 ± 1.9 | 14.4 ± 2.1 | 15.2 ± 1.4 | 0.133 |
| Creatinine (mg/dl) | 0.82 ± 0.26 | 0.85 ± 0.28 | 0.78 ± 0.22 | 0.408 |
| e-GFR (ml/min/1.73 m<sup>2</sup>) | 73.8 ± 3.0 | 71.0 ± 22.7 | 78.2 ± 17.8 | 0.246 |
| Knee extension strength<br>(Nm/kg) | 1.8 (1.5–2.5) | 1.7 (1.5–2.0) | 2.3 (1.2–2.8) | 0.285 |
| Skeletal muscle index (kg/m<sup>2</sup>) | 7.6 ± 1.2 | 7.5 ± 1.2 | 7.7 ± 1.2 | 0.526 |
| Fatigability |  |  |  |  |
| ΔMVCC velocity (%) | 35.9 ± 17.0 | 39.8 ± 17.4 | 29.5 ± 14.6 | 0.036<sup>#</sup> |
| ΔMVIC torque (%) | 26.7 ± 15.6 | 29.8 ± 14.3 | 21.5 ± 16.5 | 0.067 |
| Physical activity |  |  |  |  |
| Low [N, (%)] | 9 (18.0) | 9 (29.0) | 0 (0.0) |  |
| Moderate [N, (%)] | 26 (52.0) | 15 (48.4) | 11 (57.9) | 0.027<sup>#</sup> |
| High [N, (%)] | 15 (30.0) | 7 (22.6) | 8 (42.1) |  |

Data are expressed as numbers (percentages) or mean ± standard deviation or median (interquartile range).
The p-value indicates a significant difference between the DKD group and the non-DKD group using unpaired t test, Mann–Whitney U test, $\chi^2$ test, or Fisher’s exact test.
*BMI* body mass index, *HbA1c* hemoglobin A1c, *e-GFR* estimated glomerular filtration rate, *MVCC* maximal voluntary concentric contractions, *MVIC* maximal voluntary isometric contractions.
<sup>#</sup>p < 0.05

### Kidney disease

Of the participants in this study, 31 (62%) had concomitant DKD, and the demographic characteristics of the DKD and non-DKD groups were similar. Median knee extensor strength was 1.7 Nm/kg (IQR, 1.5–2.0) in the DKD group and 2.3 Nm/kg (IQR, 1.2–2.8) in the non-DKD group, without significant differences. Similarly, the mean (SD) SMI showed no significant difference between the DKD (7.5 kg/m^2^, 1.2) and non-DKD groups (7.7 kg/m^2^, 1.2) (Table 1).

### Fatigability

The mean (SD) ΔMVCC velocity for all participants was 35.9% (17.0), and the mean ΔMVIC torque was 26.7% (15.6). The mean ΔMVCC velocity of the DKD group was 39.8% (17.4), and that of the non-DKD group was 29.5% (14.6); the DKD group had a significantly higher ΔMVCC velocity (Table 1, Fig 3).

**Fig 3.**
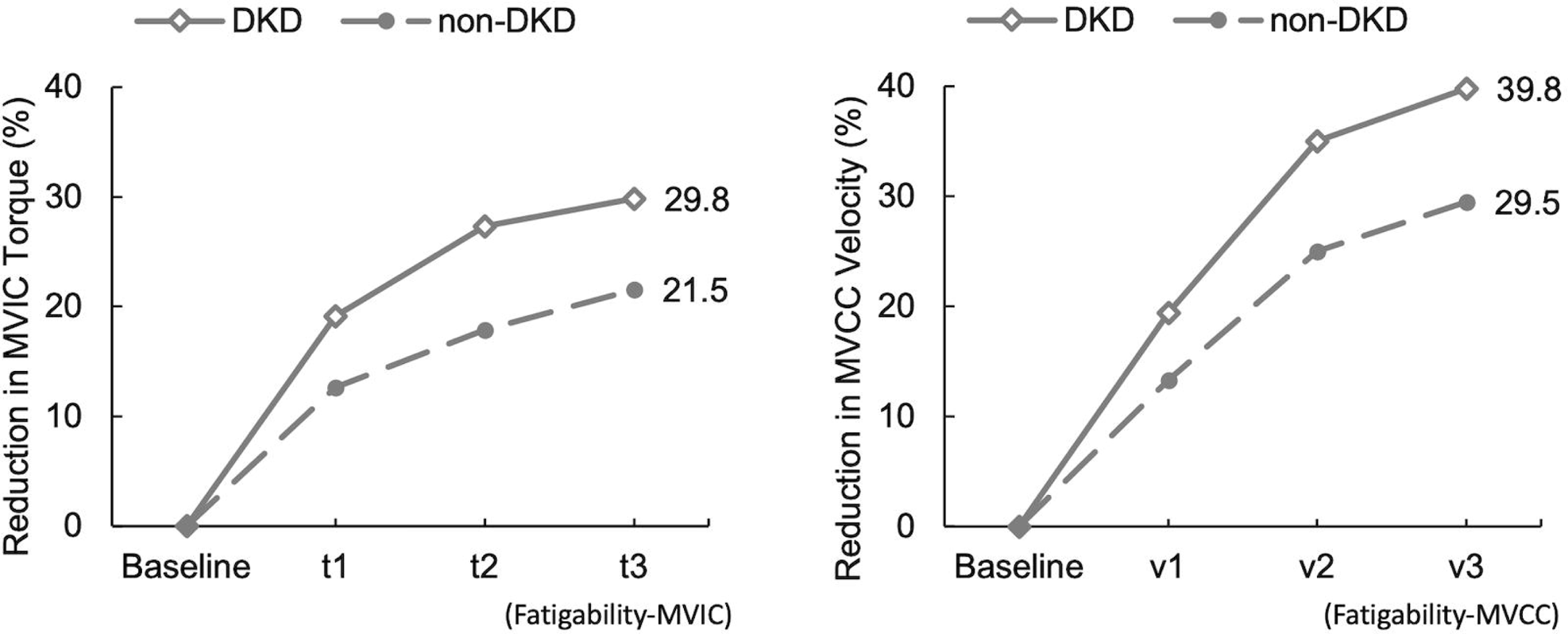
Differences in fatigability between DKD and non-DKD groups. This showed the MVCC velocity and MVIC torque during the baseline to dynamic fatiguing task. ΔMVCC velocity at t3 and ΔMVIC torque at v3 indicated fatigability in this study. *MVCC* maximal voluntary concentric contractions, *MVIC* maximal voluntary isometric contractions, *DKD* diabetic kidney disease.

### Relationship between the presence of kidney disease and fatigability

The results of the regression analysis assessing the effect of kidney disease on fatigability were: After adjusting for age, sex, knee extensor strength, and SMI, ΔMVCC velocity was significantly higher in the DKD group compared with the non-DKD group (F=4.235, p<0.05). After adjusting for similar variables, ΔMVIC torque was significantly higher in the DKD group compared with the non-DKD group (F=4.853, p<0.05) (Table 2).

**Table 2.**
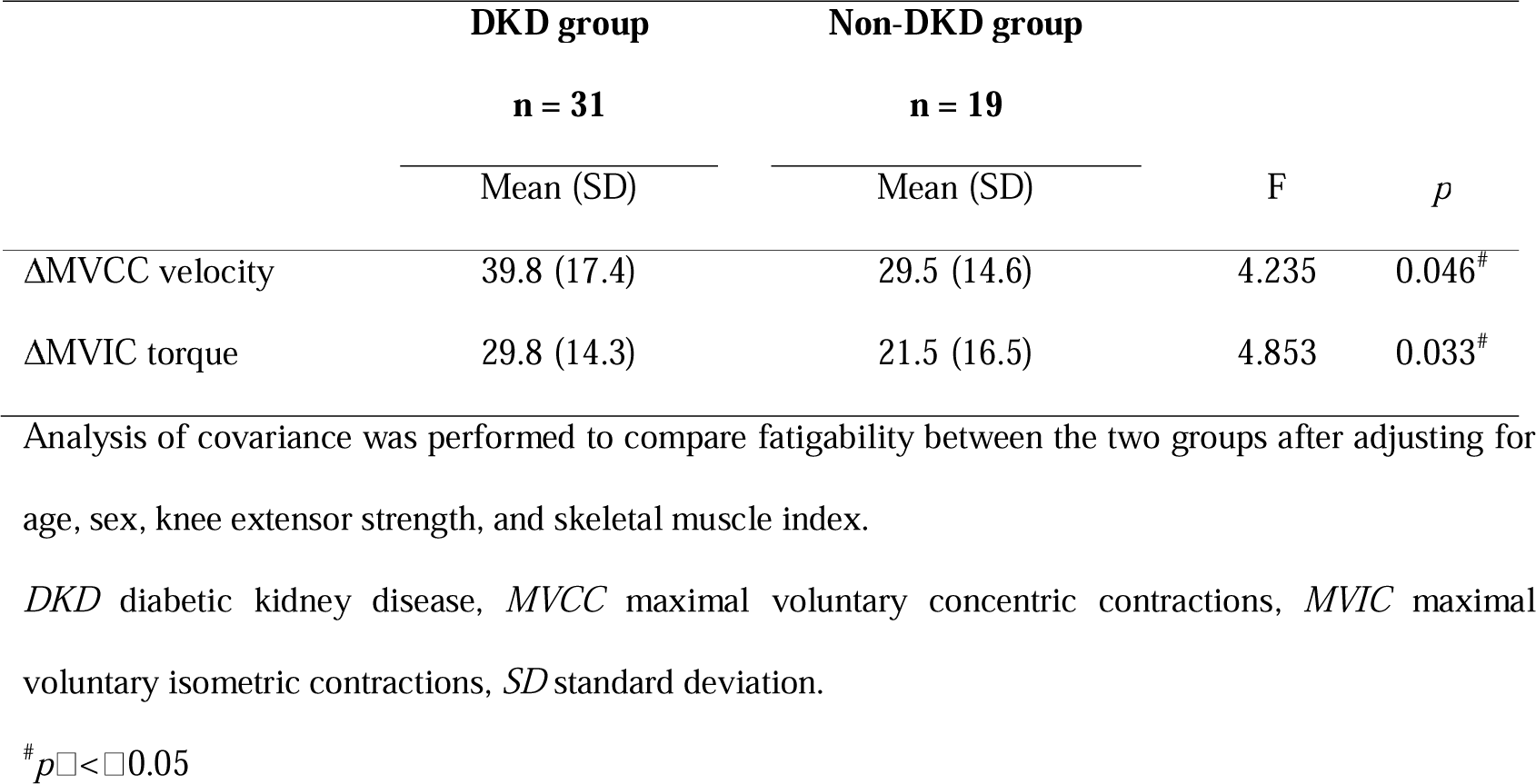
Fatigability comparison with and without DKD.

|  | DKD group | Non-DKD group |  |  |
| --- | --- | --- | --- | --- |
|  | n = 31 | n = 19 |  |  |
|  | Mean (SD) | Mean (SD) | F | p |
| ΔMVCC velocity | 39.8 (17.4) | 29.5 (14.6) | 4.235 | 0.046<sup>#</sup> |
| ΔMVIC torque | 29.8 (14.3) | 21.5 (16.5) | 4.853 | 0.033<sup>#</sup> |

Analysis of covariance was performed to compare fatigability between the two groups after adjusting for age, sex, knee extensor strength, and skeletal muscle index.
DKD diabetic kidney disease, MVCC maximal voluntary concentric contractions, MVIC maximal voluntary isometric contractions, SD standard deviation.
<sup>#</sup> $p \leq 0.05$

### Physical activity

Of all the participants, 9 (18%) had low physical activity, 26 (52%) had moderate physical activity, and 15 (30%) had high physical activity. All 9 patients with low physical activity were in the DKD group, and the DKD group had a significantly higher percentage of low physical activity than did the non-DKD group (29% vs. 0%, p<0.05). The percentages of moderate and high physical activities did not differ significantly between the DKD and non-DKD groups (Table 1).

### Relationship between fatigability and physical activity

The results of ordinal logistic regression analysis adjusted for age, sex, knee extensor strength, and SMI for all participants and the DKD and non-DKD groups were: In all participants, both ΔMVCC velocity and ΔMVIC torque were significantly associated with physical activity (odds ratio [OR], 0.952; 95% confidence interval [CI]: 0.919–0.987, p<0.05/OR, 0.962; 95% CI: 0.926–0.998, p<0.05, respectively). In the DKD group, ΔMVCC velocity was significantly associated with physical activity (OR: 0.045, 95% CI: 0.913–0.999, p<0.05). However, in the non-DKD group, neither ΔMVCC velocity nor ΔMVIC torque was associated with physical activity (Tables 3 and 4).

**Table 3.** Relationship between physical activity and ΔMVCC velocity in all participants, DKD group, and non-DKD group.

|  |  | Physical activity |  |  |  |  |  |
| --- | --- | --- | --- | --- | --- | --- | --- |
|  |  | All |  | DKD group |  | Non-DKD group |  |
|  |  | OR (95% CI) | *p*-Value | OR (95% CI) | *p*-Value | OR (95% CI) | *p*-Value |
| Sex | Female | 1 |  | 1 |  | 1 |  |
|  | Male | 1.224 (0.271–5.529) | 0.793 | 2.691 (0.321–22.511) | 0.361 | 0.785 (0.027–22.692) | 0.888 |
| Age |  | 1.041 (0.958–1.132) | 0.340 | 1.008 (0.907–1.121) | 0.878 | 1.213 (0.994–1.480) | 0.057 |
| ΔMVCC velocity | | 0.952 (0.919–0.987) | 0.007<sup>#</sup> | 0.955 (0.913–0.999) | 0.045<sup>#</sup> | 0.968 (0.881–1.063) | 0.494 |
| Knee extension strength |  | 1.103 (0.493–2.467) | 0.811 | 0.481 (0.107–2.155) | 0.339 | 1.343 (0.315–5.720) | 0.690 |
| Skeletal muscle index |  | 0.934 (0.503–1.735) | 0.830 | 0.751 (0.350–1.611) | 0.462 | 1.782 (0.433–7.323) | 0.423 |

To determine the relationship between physical activity and ΔMVCC velocity, ordinal logistic regression analysis was performed considering sex, age, knee extension strength, and skeletal muscle index in all participants, DKD group, and non-DKD group.
*DKD* diabetic kidney disease, *OR* odds ratio, *CI* confidence interval, *MVCC* maximal voluntary concentric contractions.
<sup>#</sup>*p* < 0.05

**Table 4.**
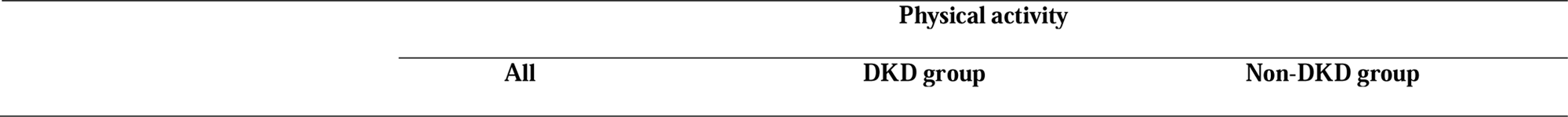
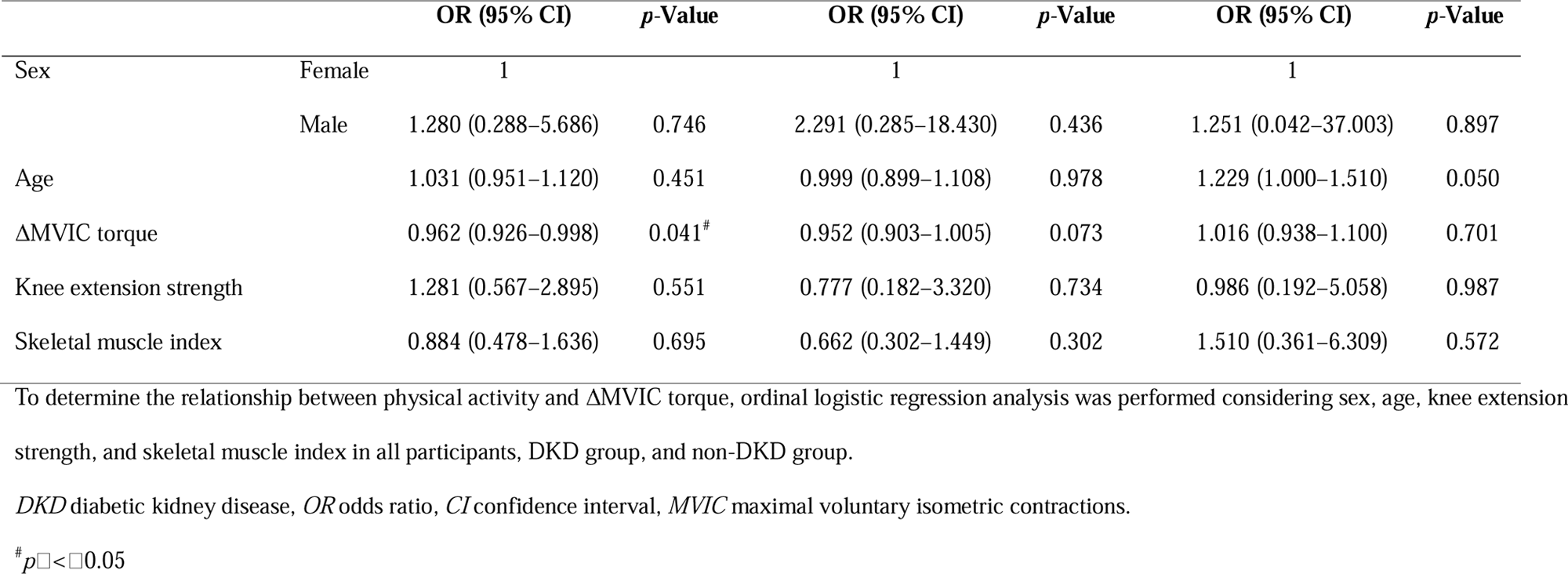
Relationship between physical activity and ΔMVIC torque in all participants, DKD group, and non-DKD group.

|  |  | Physical activity |  |  |
| --- | --- | --- | --- | --- |
|  |  | All | DKD group | Non-DKD group |

|  |  | **OR (95% CI)** | ***p*-Value** | **OR (95% CI)** | ***p*-Value** | **OR (95% CI)** | ***p*-Value** |
| --- | --- | --- | --- | --- | --- | --- | --- |
| Sex | Female | 1 |  | 1 |  | 1 |  |
|  | Male | 1.280 (0.288–5.686) | 0.746 | 2.291 (0.285–18.430) | 0.436 | 1.251 (0.042–37.003) | 0.897 |
| Age |  | 1.031 (0.951–1.120) | 0.451 | 0.999 (0.899–1.108) | 0.978 | 1.229 (1.000–1.510) | 0.050 |
| ΔMVIC torque |  | 0.962 (0.926–0.998) | 0.041<sup>#</sup> | 0.952 (0.903–1.005) | 0.073 | 1.016 (0.938–1.100) | 0.701 |
| Knee extension strength |  | 1.281 (0.567–2.895) | 0.551 | 0.777 (0.182–3.320) | 0.734 | 0.986 (0.192–5.058) | 0.987 |
| Skeletal muscle index |  | 0.884 (0.478–1.636) | 0.695 | 0.662 (0.302–1.449) | 0.302 | 1.510 (0.361–6.309) | 0.572 |

To determine the relationship between physical activity and ΔMVIC torque, ordinal logistic regression analysis was performed considering sex, age, knee extension strength, and skeletal muscle index in all participants, DKD group, and non-DKD group.
DKD diabetic kidney disease, OR odds ratio, CI confidence interval, MVIC maximal voluntary isometric contractions.
<sup>#</sup> $p < 0.05$

## Discussion

Fatigability was greater among patients with T2DM combined with DKD compared with those with T2DM providing evidence that DKD is associated with greater fatigability among patients with T2DM. Furthermore, fatigability may be related to the amount of physical activity required by patients with DKD. Facilitating increased physical activity is crucial for individuals with DM, contributing to glycemic control, preventing diabetic complications, and reducing the incidence of cardiovascular disease and mortality [22,23]. A novel aspect of our study is that the focus on fatigability was increased by DKD complications, suggesting that this increased fatigability may be associated with decreased physical activity. Unlike chronic fatigue, fatigability can be modified with exercise therapy [24]. Therefore, this study is clinically significant because it may identify a therapeutic target to increase physical activity. In a previous study reporting fatigability in healthy older adults averaging approximately 70 years of age, ΔMVCC velocity (male/female) was approximately 24.7%/23.1%, and ΔMVIC torque (male/female) was approximately 23.2%/17.1% [16]. However, in this study involving patients aged 40–65 years, ΔMVCC velocity was 29.5% and 39.8% in patients with non-DKD and DKD, respectively, whereas ΔMVIC torque was 21.5% and 29.8% in patients with non-DKD and DKD, respectively. In other words, even in patients with DM, fatigability rises to a level comparable to that in healthy elderly individuals. Furthermore, when combined with DKD and diabetes, fatigability rises by more than 1.5 times that of healthy elderly individuals, indicating its potential as a significant factor inhibiting an increase in physical activity.

### Mechanism of fatigability and involvement of DKD complications in fatigability

The following biological mechanisms are considered responsible for increased fatigability, including activation of the motor cortex, descending motor neurons, spinal activation of motor neurons, neuromuscular junction transmission, muscle perfusion (blood flow), muscle fiber excitation-contraction, and energy metabolism (adenosine triphosphate regulation), and impairment of this process increases fatigability [25]. A study comparing the fatigability of healthy adults and T2DM reported increased fatigability in T2DM due to muscle contractile properties [26]. Substitution of the general process suggests that excitation-contraction in muscle fibers is impaired. Besides this mechanism, increased oxidative stress, skeletal muscle abnormalities due to CKD complications, and impaired microcirculation due to decreased nitric oxide production may be factors that further increase fatigability in the DKD group. Microcirculatory disturbances due to CKD complications reduce the rate of oxygen uptake into tissues and impair oxygen delivery to mitochondria in muscle tissue [27,28]. Impaired oxygen delivery to muscle tissue reduces adenosine triphosphate utilization efficiency and decreases calcium ion sensitivity in myofibrils and calcium ion release capacity in the sarcoplasmic reticulum [11]. This phenomenon occurs even in patients with mild renal impairment [29]. Furthermore, trace albuminuria is associated with endothelial dysfunction, sarcopenia, and muscle strength [30–32], suggesting that peripheral tissue abnormalities may also affect fatigability in early-stage patients, such as our participants. In other words, the complications of DKD contribute to the process of increased fatigability in T2DM and enhance fatigability by inducing biological changes in peripheral tissues due to the complications of CKD.

### Relationship between fatigability and physical activity

Studies on healthy adults have reported a relationship between fatigability and physical activity [16,33](. However, the association remains unknown in the T2DM and DKD. The brain activity that occurs during the preparatory phase of physical activity (motor-related cortical potential) is associated with the perception of effort [34]. Moreover, greater muscle fatigability increases motor-related cortical potential [35]. In other words, patients with increased fatigability have greater perceived effort during physical activity. Increases in perceived effort decrease willingness to initiate physical activity [36]. Additionally, patients with increased fatigability require more effort for cognitive processing because of reduced oxygen supply to the brain due to excessive muscle hypermetabolism during physical activity [36]. Mild renal impairment contributes to this process because of poor oxygen supply to the brain during activity [37]. Thus, increased fatigability may be a barrier to the initiation and continuation of physical activity, which may be further increased by the complications of DKD.

### Limitations

This study has some limitations. First, the sample size was small owing to the single-center nature of the study, and the fact that it was conducted with a limited number of subjects to account for age effects reduced the generalizability and external validity. Second, the IPAQ-SF relies on subjective self-reporting, which can be affected by respondents’ perceptions and memory biases, potentially leading to inaccuracies, particularly in individuals with fluctuating activity levels or vague recall of specific activities. Third, we did not consider the impact of diabetic neuropathy (DN) as a factor related to fatigue. Currently, the gold standard for DN is nerve conduction testing, which was not performed in this study. However, knee extension strength and skeletal muscle mass were adjusted for as confounding factors in this study. Muscle strength has been reported to be associated with the presence and severity of DN, although not with albuminuria or retinopathy itself [38]. Moreover, patients with DN have been reported to have a greater annual loss of muscle mass in the lower extremities than patients without DN, confirming the association between DN and skeletal muscle mass [39]. Therefore, in this study, the effect of DN on fatigability was minimal. Finally, the possibility that the severity and duration of DM itself are factors contributing to increased fatigability was not taken into account. DM progression is associated with DKD complications and increased fatigability. However, adjusting for this adaptive confounding factor was not possible due to the lack of standardization in the severity assessment of T2DM, and the clinical assessment of DM duration is challenging.

## Conclusions

This study showed that complications of DKD in patients with T2DM increased activity-induced fatigability. The findings also suggested a potential correlation between fatigability and the level of physical activity, with a similar trend observed specifically among patients with DKD. These results aim to shed light on factors associated with physical activity levels in patients with DKD and contribute to the development of tailored physical activity and exercise programs based on patient characteristics. In the future, conducting longitudinal studies to verify the effect of reducing fatigue on increasing physical activity is crucial.

## Data Availability

Data presented in this study are available upon request from the corresponding author. The data are not publicly available because they are the property of the Institute of Hamamatsu University Hospital, Japan. Data are available from the Hmamatsu University School of Medicine Data Access (contact via Yuma Hirano) for researchers who meet the criteria for access to confidential data.

## Acknowledgments

The authors thank the patients at the Hamamatsu University Hospital who participated in this study. The authors would like to thank the staff and nurses for their excellent patient care. We would like to thank Editage (www.editage.com) for the English language editing.

